# Safety of the NVX-CoV2373 COVID-19 Vaccine in Randomized Placebo-Controlled Clinical Trials

**DOI:** 10.1101/2023.02.24.23285601

**Authors:** Katherine Smith, Karim Hegazy, Miranda R. Cai, Irene McKnight, Matthew D. Rousculp, Katia Alves

## Abstract

**Background:** NVX-CoV2373 (Nuvaxovid™ or the Novavax COVID-19 Vaccine, Adjuvanted), the first protein-based COVID-19 vaccine, received emergency use authorization (EUA) as a primary series/booster and is available globally. NVX-CoV2373 primary series demonstrated efficacy rates of 89·7–90·4 % and an acceptable safety profile. This article summarizes safety in adult recipients (aged ≥18 years) of primary series NVX-CoV2373 in four randomized placebo-controlled trials.

**Methods:** All participants who received NVX-CoV2373 primary series or placebo (pre-crossover) were included according to actual received treatment. The safety period was from Day 0 (first vaccination) to unblinding/receipt of EUA-approved/crossover vaccine, end of each study (EOS), or last visit date/cutoff date minus 14 days. The analysis reviewed local and systemic solicited adverse events (AEs) within 7 days after NVX-CoV2373 or placebo; unsolicited AEs from after Dose 1 to 28 days after Dose 2; serious AEs (SAEs), deaths, AEs of special interest, and vaccine-related medically attended AEs from Day 0 through end of follow-up (incidence rate per 100 person-years).

**Findings:** Pooled data from 49,950 participants (NVX-CoV2373, n=30,058; placebo, n=19,892) were included. Solicited reactions after any dose were more frequent in NVX-CoV2373 recipients (local, 76%/systemic, 70%) than placebo recipients (29%/47%), and were mostly of mild-to-moderate severity. Grade 3+ reactions were infrequent, with greater frequency in NVX-CoV2373 recipients (6·28%/11·36%) than placebo recipients (0·48%/3·58%). SAEs and deaths occurred with similarly low frequency in NVX-CoV2373 (0·91% and 0·07%, respectively) and placebo recipients (1·0% and 0·06%).

**Interpretation:** To date, NVX-CoV2373 has displayed an acceptable safety profile in healthy adults.

**Funding:** Supported by Novavax, Inc.

## Introduction

The novel human coronavirus disease 2019 (COVID-19) was first reported in 2019, and subsequently spread globally to become the fifth pandemic documented since the flu pandemic in 1918.^1^ To date, over 6·5 million people have died from the disease.^2^ People who are fully vaccinated but experience a breakthrough infection are less likely to have serious COVID-19 symptoms than those who are unvaccinated.^3^ COVID-19 vaccines (primary series and boosters) have been shown to be effective at protecting against serious illness, hospitalization, and death.^4^ NVX-CoV2373 (Nuvaxovid™ or the Novavax COVID-19 Vaccine, Adjuvanted), the first protein-based COVID-19 vaccine, recently received emergency use authorization from the FDA^5^ and is available in most countries around the world.^6^ The vaccine has been shown to be effective and safe in clinical trials conducted to date.^7–11^

Here, we present the results of a summary of safety in 49,950 adults aged ≥18 years who received at least one dose of NVX-CoV2373 or placebo across the clinical development program, using pooled data from the four clinical trials in which the primary endpoint analysis has been completed.

## Methods

**Figure 1** and **Table S1** summarize the key design features of the clinical trials evaluating NVX-CoV2373 that were included in the safety analysis for four studies. The four studies selected represent all trials in which primary endpoint analyses were completed at the time this safety summary was performed. For the purpose of this safety analysis, all participants who received at least one dose of NVX-CoV2373 or placebo were included. The phase 1/2 2019nCoV-101 study (US and Australia; NCT04368988) evaluated multiple formulations of the vaccine and Matrix-M™ adjuvant; thus, only the participants in Treatment Groups A and C in Part 1 and participants in Treatment Groups A, B, and C in Part 2 (participants receiving the approved 5 ug dose) are included in the analysis. The phase 2A/B 2019nCoV-501 study (South Africa; NCT04533399) assessed vaccine efficacy, safety, and immunogenicity in participants aged 18–84 years, with and without underlying HIV-1. Studies 2019nCoV-301 (US and Mexico; NCT04611802) and 2019nCoV-302 (UK; EudraCT number, 2020-004123-16) are efficacy studies evaluating the durability of vaccine efficacy in adult participants aged 18–84 years. These two studies included a non–placebo-controlled crossover period where participants who received placebo were given NVX-CoV2373; however, the analysis described here was performed using only the data collected during the pre-crossover period. Novavax, Inc. funded and conducted the trials discussed in this analysis and funded preparation of the manuscript.

**Figure 1:**
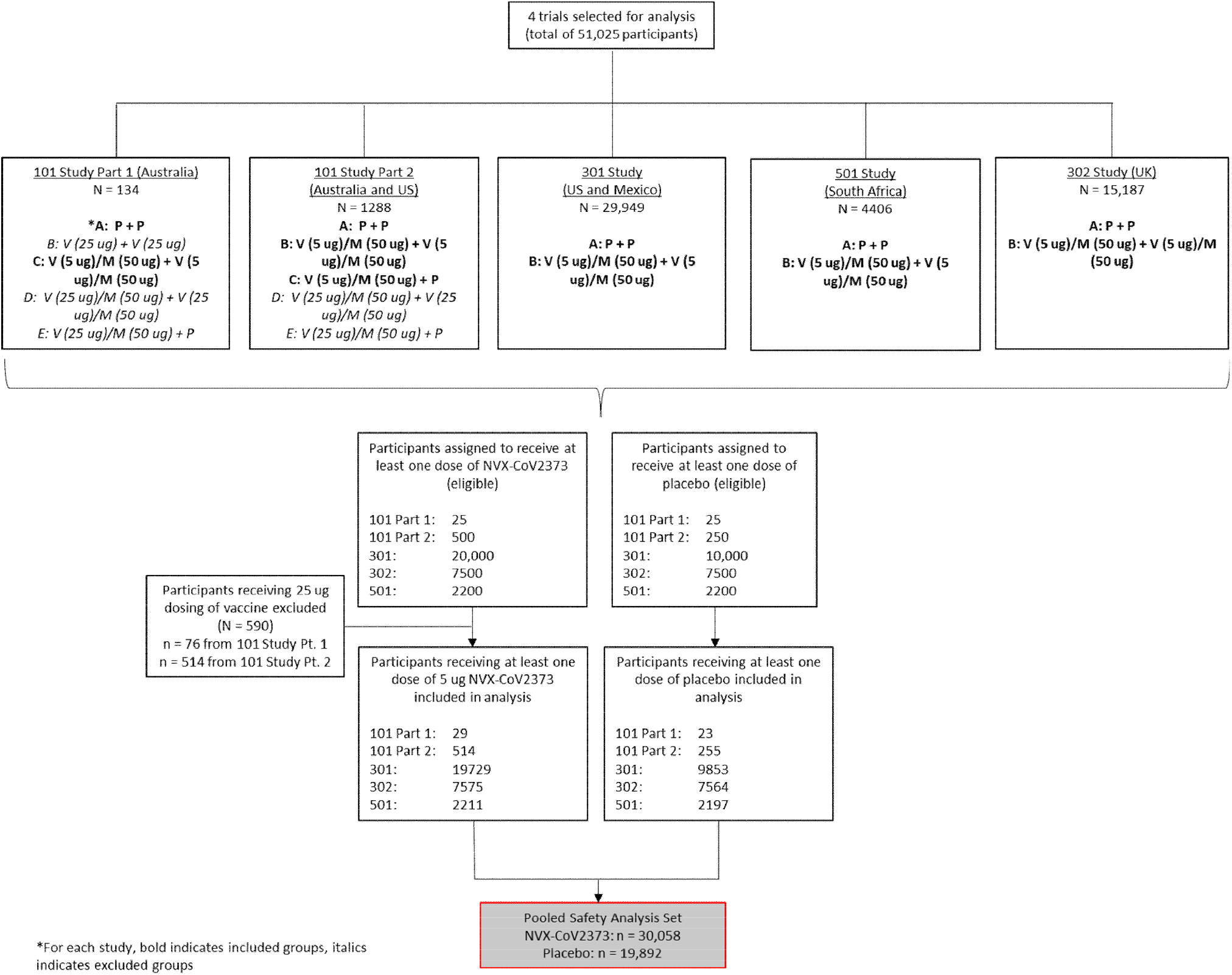
Study inclusion diagram. Four trials were completed at the time of this analysis and were selected for inclusion. Safety data on participants from each trial (Study 101 had two phases) were pooled. Only participants who received the approved 5 ug dose of NVX-CoV2373 or placebo were included. Abbreviations: M, Matrix-M™ adjuvant; P, placebo; V, active NVX-CoV2373 vaccine.

### Study population

The safety analysis set included all participants who received at least one dose of 5 ug NVX-CoV2373 or placebo (**Figure 1**). Participants were included in the analysis according to the study vaccine (NVX-CoV2373 or placebo) received. For participants who receive a mixed regimen of the study vaccine, their safety data were analyzed based on the following:

1. If active vaccine was given at Dose 1 and placebo was given at Dose 2: participants were included in the vaccine group but not included in the post-Dose 2 short-term AE summary (ie, within 28 days after Dose 2)
2. If placebo was given at Dose 1 and active vaccine was given at Dose 2: participants were included in the vaccine group with safety follow-up starting from Dose 2

### Safety analyses

Safety data are reported for all participants who received at least one dose of vaccine or placebo. Solicited adverse events (AEs; local and systemic reactions) reported within 7 days after the administration of NVX-CoV2373 or placebo were assessed. Unsolicited AEs reported from Day 0 to 49 (28 days post-Dose 2), split by age group, were also assessed. Serious AEs (SAEs) and deaths throughout the follow-up period were identified. Additionally, AEs of special interest (AESIs), focused on potential immune-mediated medical conditions (PIMMCs) based on the definitions given in the FDA guidelines, were collected throughout the follow-up period. Vaccine-related medically attended AEs (MAAEs) were collected throughout the studies. Due to different follow-up times of participants in the trials, the event rates of SAEs, deaths, and AESIs (including PIMMCs) were adjusted for the follow-up time and presented as incidence rates per 100 person-years (PY). An analysis was also done to look at cases of myocarditis/pericarditis.

Safety events were summarized descriptively. AEs were coded by preferred term and system organ class using Version 23·1 of the Medical Dictionary for Regulatory Activities (MedDRA) and summarized by severity and relationship to study vaccine. For participants who enrolled in the blinded crossover, safety data were censored at the time of crossover (ie, the date at which participants received their third study dose). For participants who did not enter the blinded crossover and were unblinded (but did not withdraw from the study), safety data were censored at the time of unblinding or the time of receipt of another COVID-19 vaccine (whichever date was noted first). For participants who neither entered the blinded crossover nor were censored, the safety data cutoff date was 15 March 2022.

## Statistical analysis

### Safety analysis

Summaries of safety parameters (count and percent or event rate per 100 PY) are provided by treatment group and by severity as appropriate (reported for all participants and for age groups). If a two-sided 95% confidence interval (CI) was required for the proportion of AEs, the Clopper Pearson exact method was used.^12^ This CI was calculated using SAS PROC FREQ with the exact option for binomial proportions.

For summaries of long-term AEs where trial participants had different lengths of follow-up, the incidence rate of AEs was adjusted for the length of follow-up time and calculated as incidence per 100 PY. If a 95% CI was required for the incidence rate of AEs, the exact method based on the Poisson distribution was used.^13^

Risk differences (vaccine – placebo) are provided for AESIs. The risk difference was calculated as a weighted average of risk differences across individual studies, using Cochran-Mantel-Haenszel (CMH) weights based on the sample sizes of individual studies.^14,15^ If statistical comparisons and/or 95% CIs for risk differences were required, the CMH method was used to calculate P values and CIs. The analysis was stratified by study to account for different randomization ratios and other potential differences in individual studies.

If the number of events within a stratum was too low (eg, <5) and/or the computation became unstable due to small cell counts, the sparse strata were combined with other strata or the analysis was performed without stratification. In the extreme case when the CMH method could not be implemented, the CI was computed using the method of Miettinen and Nurminen (1985).^16^ The CMH method was implemented using SAS procedures.

## Results

### Included studies and participants

Four different studies were included in the pooled safety analysis: 2019nCoV-101 Part 1 (Australia) and Part 2 (Australia, US), 2019nCoV-501 (South Africa), 2019nCoV-302 (United Kingdom), and 2019nCoV-301 (US, Mexico). The combined analysis included a total of 49,950 participants—30,058 participants who received vaccine, and 19,892 who received placebo (**Table S1, Figure 1**).

For all included participants, mean (SD) age was 47·4 (15·88) years (47·2 [15·78] years for NVX-CoV2373, 47·7 [16·03] years for placebo), with approximately 83–84% of participants in the 18–64 years group. The participants were similarly distributed between sexes (approximately 48% female, 52% male) and were primarily White (approximately 75%) (**Table 1**).

**Table 1:** Pooled demographic data from analyzed trials.

| Parameter | NVX-CoV2373 (n=30,058) | Placebo (n=19,892) | Total (n=49,950) |
| --- | --- | --- | --- |
| Age, mean (SD) years | 47·2 (15·78) | 47·7 (16·03) | 47·4 (15·88) |
| 18–64 years, n (%) | 25282 (84·11) | 16433 (82·61) | 41715 (83·51) |
| 65 years and older, n (%) | 4776 (15·89) | 3459 (17·39) | 8235 (16·49) |
| Sex |  |  |  |
| Female, n (%) | 14232 (47·35) | 9528 (47·90) | 23760 (47·57) |
| Male, n (%) | 15826 (52·65) | 10364 (52·10) | 26190 (52·43) |
| Race, n (%) |  |  |  |
| White | 22415 (74·57) | 14808 (74·44) | 37223 (74·52) |
| Black or African American | 4417 (14·69) | 3256 (16·37) | 7673 (15·36) |
| Asian | 1119 (3·72) | 691 (3·47) | 1810 (3·62) |
| American Indian or Alaska Native | 1322 (4·40) | 665 (3·34) | 1987 (3·98) |
| Native Hawaiian or Other Pacific Islander | 58 (0·19) | 13 (0·07) | 71 (0·14) |
| Multiple | 463 (1·54) | 260 (1·31) | 723 (1·45) |
| Not reported | 209 (0·70) | 134 (0·67) | 343 (0·69) |
| Other | 43 (0·14) | 54 (0·27) | 97 (0·19) |
| Missing | 12 (0·04) | 11 (0·06) | 23 (0·05) |
| Hispanic or Latino, n (%) | 4463 (14·85) | 2262 (11·37) | 6725 (13·46) |

SD=standard deviation.

### Safety

#### Adverse events summary

Approximately 76% of participants who received NVX-CoV2373 and 29% of participants receiving placebo experienced solicited local reactions within 7 days. The frequency of these events was higher for Dose 2 than Dose 1 for NVX-CoV2373 only; the placebo group had a lower frequency of local reactions at Dose 2 than Dose 1. Grade 3+ reactions were infrequent (6·28% for NVX-CoV2373, 0·48% for placebo). Approximately 70% of NVX-CoV2373 recipients and 47% of placebo recipients had solicited systemic reactions within 7 days. The frequency of these reactions was higher for Dose 2 than Dose 1 in the vaccine group only. Again, Grade 3+ reactions were infrequent (11·36% for NVX-CoV2373, 3·58% for placebo). For both local and systemic reactions (overall and Grade 3+), the NVX-CoV2373 group had higher frequencies of reactions than the placebo group regardless of dose (**Table 2**).

**Table 2:** Summary of all adverse event types reported during the study (Day 0 to end of follow-up)

| Event, subject n (%) | NVX-CoV2373 | Placebo |
| --- | --- | --- |
| **Reactogenicity** | **N=27,797** | **N=13,004** |
| Solicited injection site reaction within 7 days | 17,333 (76.03%) | 3782 (29.08%) |
| Dose 1/Dose 2 | 12,178 (53.42%) / 15,213 (66.73%) | 2510 (19.30%) / 2241 (17.23%) |
| Grade 3+ (any dose) | 1432 (6.28%) | 62 (0.48%) |
| Solicited systemic reaction within 7 days | 15,870 (69.61%) | 6141 (47.22%) |
| Dose 1/Dose 2 | 10,077 (44.20%) / 13,252 (58.13%) | 4675 (35.95%) / 3767 (28.97%) |
| Grade 3+ (any dose) | 2589 (11.36%) | 465 (3.58%) |
| **Other AEs** | **N=30,077** | **N=19,871** |
| Any unsolicited AEs | 6266 (20.83%) | 3656 (18.40%) |
| Non-serious unsolicited AE | 6102 (20.29%) | 3539 (17.81%) |
| *Treatment-related nonserious unsolicited AEs* | 2713 (9.02%) | 924 (4.65%) |
| Grade 3 non-serious unsolicited AE | 220 (0.73%) | 109 (0.55%) |
| *Treatment-related Grade 3 non-serious unsolicited AE* | 81 (0.27%) | 17 (0.09%) |
| Any SAEs | 275 (0.91%) | 198 (1.00%) |
| Treatment-related SAEs | 8 (0.03%) | 4 (0.02%) |
| MAAEs | 1598 (5.31%) | 966 (4.86%) |
| Treatment-related MAAEs | 146 (0.49%) | 52 (0.26%) |
| AEs leading to discontinuation | 82 (0.27%) | 36 (0.18%) |
| PIMMCs (CRF or SMQ) | 45 (0.15%) | 31 (0.16%) |
| Deaths | 21 (0.07%) | 12 (0.06%) |

AE=adverse event; CRF=case report form; MAAE=medically attended adverse event; PIMMC=potential immune-mediated medical condition; SAE=serious adverse event; SMQ=standardized MedDRA query.

The frequency of overall unsolicited AEs was similar between the NVX-CoV2373 and placebo groups (20·83% and 18·40%, respectively), as was the frequency of Grade 3+ reactions (0·73% and 0·55% percent, respectively). Treatment-related unsolicited AEs were slightly higher for NVX-CoV2373 than placebo (overall: 9·02% vs. 4·65%; Grade 3+: 0·27% vs. 0·09%). SAEs, MAAEs, and AEs leading to discontinuation all occurred at similar frequencies between the NVX-CoV2373 and placebo groups. PIMMCs and deaths were rare in both groups: 0·15% vs. 0·16% for PIMMCs, and 0·07% vs. 0·06% for deaths (**Table 2**).

#### Reactogenicity (solicited reactions)

Solicited local reactions were higher in the NVX-CoV2373 group than in the placebo group; however, few were classified as Grade 3 or 4 (**Table S2**). The most frequent local reaction was pain/tenderness (**Figure 2A**). Solicited systemic events also appeared at a higher rate in the NVX-CoV2373 group: almost twice as high for Dose 2 as in the placebo group (**Table S3**). The most frequent systemic reactions were fatigue/malaise, headache, and muscle pain (**Figure 2B**). Solicited reactions were mostly mild and generally resolved quickly (within 7 days). The most common reactions that persisted beyond 7 days were pain/tenderness, headache, and fatigue/malaise (**Table S4**). When the rates of solicited AEs were stratified by age, the rates of local and systemic events were higher in the 18–64 years group than in the >65 years group, after both Dose 1 and Dose 2 (**Tables S2, S3**). Notably, fever occurred in less than 1% of participants following Dose 1 and 6% of participants following Dose 2 (**Table S3**).

**Figure 2:**
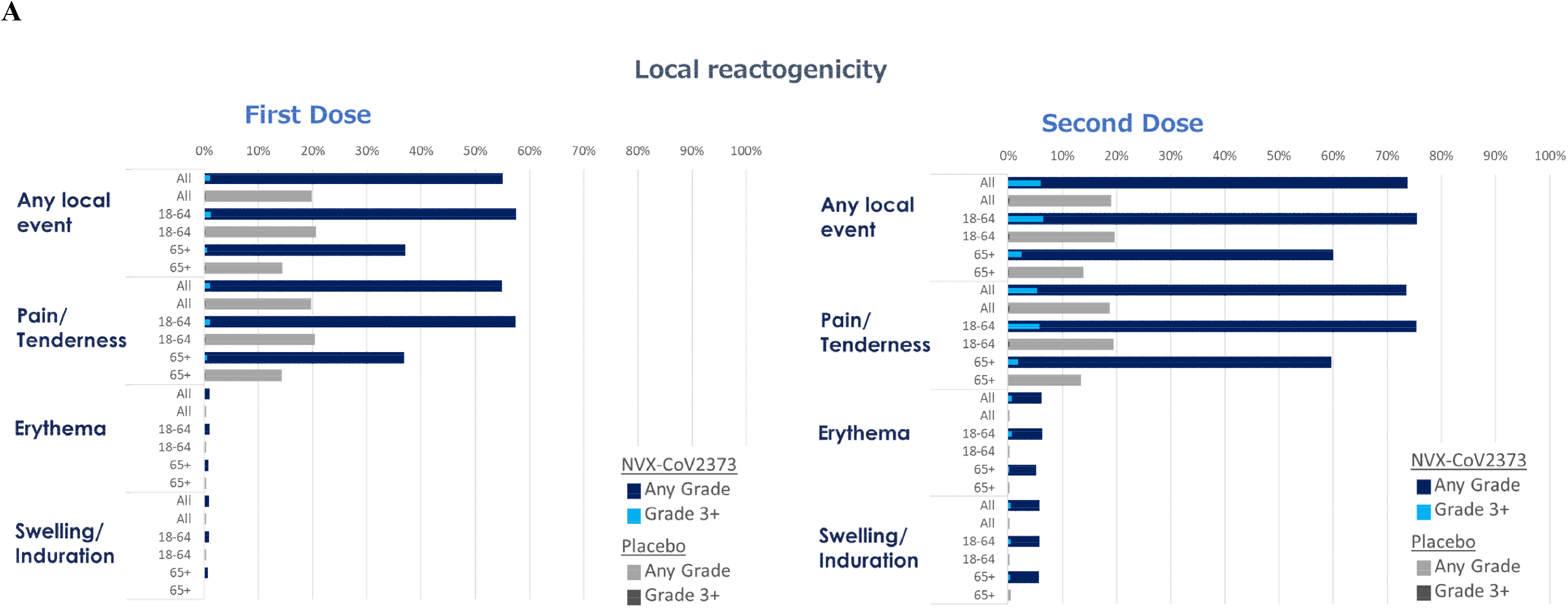
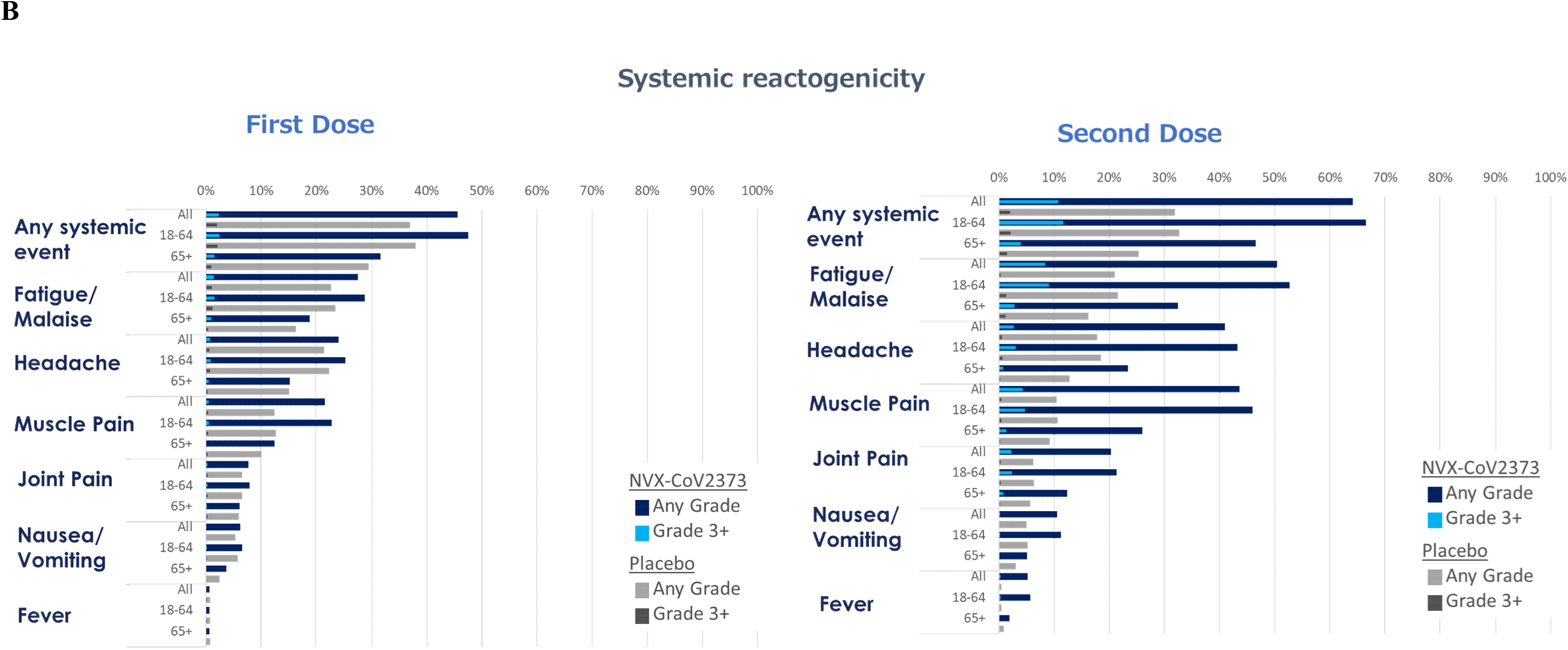
Local and systemic reactogenicity. Solicited local (A) and systemic (B) reactions are shown for Dose 1 and Dose 2, for all participants in each treatment group or split by age group (18–64 years and 65+ years). Frequency of reactions of any dose is plotted, as well as frequency of Grade 3+ reactions.

#### Unsolicited adverse events

The most common unsolicited AEs reported were general disorders and administration site conditions (7·21% for NVX-CoV2373, 3·37% for placebo), nervous system disorders (4·51% vs. 4·09%), musculoskeletal and connective tissue disorders (4·37% vs. 2·50%), and infections and infestations (2·96% vs. 3·41%) (**Table S5**).

Of particular interest were unsolicited AEs related to allergy or abnormal/reduced sensations, which have been reported in post-authorization use.^17^ Hypoaesthesia occurred in 0·06% of NVX-CoV2373 recipients and 0·06% of placebo recipients (risk difference [95% CI], 0 [-0·05, 0·04]). Paraesthesia occurred in 0·10% of NVX-CoV2373 recipients and 0·14% of placebo recipients (risk difference [95% CI], -0·02 [-0·09, 0·04]). Allergy to vaccine or excipient occurred in <0·01% of NVX-CoV2373 recipients and did not occur for placebo.

Rates of unsolicited SAEs were lower for the 18–64 years group than the ≥65 years group (**Table S6**). The most common SAEs by system organ class (SOC) were infections and infestations (ER 0·58 for NVX-CoV2373, 0·94 for placebo), especially those related to COVID-19; cardiac disorders (ER 0·41 vs. 0·33); and injury, poisoning, and procedural complications (ER 0·47 vs. 0·42) (**Table S6**).

AEs led to discontinuation for 0·27% of NVX-CoV2373 participants and 0·18% of placebo recipients. Among participants aged 18–64 years, the most common AEs causing discontinuation (by ER per 100 PY in either group) included cardiac disorders (ER 0·076 for NVX-CoV2373, 0·091 for placebo); general disorders and administration site conditions (ER 0·127 vs. 0·018); and infections and infestations (ER 0·127 vs. 0·073). Among participants aged 65 years or older, the most common AEs causing discontinuation included cardiac disorders (ER 0·303 vs. 0·429); general disorders and administration site conditions (ER 0·455 vs. 0·107); infections and infestations (ER 0·076 vs. 0·322); and neoplasms - benign, malignant, and unspecified (ER 0·076 vs. 0·322) (**Table S7**).

#### PIMMCs

Rates of PIMMCs for NVX-CoV2373 recipients were <1 per 100 PY across both age cohorts (0·55 events per 100 PY for all participants) and were similar to rates reported by participants who received placebo (0·43 events per 100 PY). PIMMCs of any type were seen in 0·57 per 100 PY of participants aged 18–64 years in the NVX-CoV2373 group and 0·39 per 100 PY in the placebo group. For participants ≥65 years of age, PIMMCs occurred in 0·44 per 100 PY of participants in the NVX-CoV2373 group and 0·62 per 100 PY in the placebo group. The most common PIMMCs were nervous system disorders and musculoskeletal and connective tissue disorders (**Table S8**).

#### Myocarditis and pericarditis

For reporting of myocarditis/pericarditis, both pre-crossover and post-crossover data were examined to evaluate all possible events. Two events of myocarditis were reported in the NVX-CoV2373 group, and one event was reported in the placebo group during the pre-crossover period, with a risk difference of 0 (-0·02, 0·02). Post-crossover, two participants reported two events of pericarditis and one event of myocarditis for NVX-CoV2373, and one event of myocarditis for placebo were reported, with a risk difference of 0 (-0·02, 0·05) for myocarditis and 0·02 (0·00, 0·08) for pericarditis (**Table 3**).

**Table 3:** Summary of unsolicited adverse events for myocarditis or pericarditis (SMQ) Reported throughout the studies (pre-crossover and post-crossover)

| System organ class preferred term | NVX-CoV2373 | Placebo | Risk difference (NVX-CoV2373-placebo) |
| --- | --- | --- | --- |
| Myocarditis or pericarditis (SMQ)* | n (%), (95% CI) | n (%), (95% CI) | % (95% CI) |
| Pre-Crossover Participants | N=30072 | N=19877 |  |
| Any SMQs | 2 (<0·01), (0·00, 0·02) | 1 (<0·01), (0·00, 0·03) | 0·00 (-0·02, 0·02) |
| Cardiac disorders | 2 (<0·01), (0·00, 0·02) | 1 (<0·01), (0·00, 0·03) | 0·00 (-0·02, 0·02) |
| Myocarditis | 2 (<0·01) | 1 (<0·01) | 0·00 (-0·02, 0·02) |
| Post-Crossover Participants | N=9635 | N=18,581 |  |
| Any SMQs | 2 (0·02), (0·00, 0·07) | 1 (<0·01), (0·00, 0·03) | 0·02 (-0·01, 0·07) |
| Cardiac disorders | 2<sup>†</sup> (0·02), (0·00, 0·07) | 1 (<0·01), (0·00, 0·03) | 0·02 (-0·01, 0·07) |
| Myocarditis | 1 (0·01) | 1 (<0·01) | 0·00 (-0·02, 0·05) |
| Pericarditis | 2 (0·02) | 0 | 0·02 (0·00, 0·08) |

CI=confidence interval; SMQ=standardized MedDRA query.
\*Within-group CI is computed using Clopper-Pearson. CI for Risk Difference is based on Miettinen-Nurminen. Percentages are calculated based on number of subjects in Safety Analysis set. For myocarditis/pericarditis: SMQ version: 25·0. MedDRA version: 24·0 (2019nCoV-101 Part 1, 2019nCoV-101 Part 2, and 2019nCoV-301), 23·1 (2019nCoV-302), and 23·0 (2019nCoV-501). Included study data from 2019nCoV-101 Part1 as of 28JUL2021, 2019nCoV-101 Part2 as of 11JUN2021, 2019nCoV-302 as of 27JUL2021, 2019nCoV-501 as of 27OCT2021, and 2019nCoV-301 as of 17FEB2022.
<sup>†</sup>Note that 3 events occurred in 2 participants.

## Discussion

To date, the NVX-CoV2373 COVID-19 vaccine has shown an acceptable safety profile across numerous clinical trials and in post-authorization use. As demonstrated in this summary of safety based on data from four randomized placebo-controlled clinical trials (one with two parts), local and systemic reactogenicity events (solicited local and systemic AEs) were mostly mild and transient, with <1% Grade 3–4 events. SAE rates reported by participants who received NVX-CoV2373 or placebo were similar. Rates of discontinuation due to AEs and deaths during the trials were very low and similar for both NVX-CoV2373 and placebo. PIMMCs were also infrequent and balanced between groups.

COVID-19 was more frequent for participants in the younger placebo group compared with the older age group. This was likely due to greater precautions being taken and fewer work and social responsibilities being required of participants aged ≥65 years.

There were a small number of cases of myocarditis/pericarditis observed in the trials (pre-crossover: two cases of myocarditis for NVX-CoV2373, one for placebo; post-crossover: three events in two participants, including two cases of pericarditis/one case of myocarditis for NVX-CoV2373 and one case of myocarditis for placebo). One event in the NVX-CoV2373 pre-crossover group occurred following the first dose of vaccine in a participant in the 66–70 years age range who also had COVID-19. The second event in the pre-crossover group occurred in a participant in the 16–20 years age range following the second dose. In the post-crossover period, one event occurred concurrently with strep throat, and the remaining two events were also confounded by concurrent infections. Pericarditis/myocarditis has been observed as an SAE in the mRNA (Pfizer and Moderna), viral vector (Janssen), and protein subunit (NVX-CoV2373) vaccines. Following marketing authorization of NVX-CoV2373, reports of myocarditis and pericarditis were received from several regions, including Australia and the European Union.

Based on a review of the cumulative post-authorization reports, myocarditis and pericarditis are now considered identified risks for NVX-CoV2373. This SAE is closely monitored for all COVID-19 vaccines by the FDA and CDC^18^ and by all vaccine developers that have reported cases of myocarditis and pericarditis during controlled clinical trials or post-EUA. It is unclear what specific mechanism causes the cardiac tissue inflammation. As a commonality, myocarditis related to COVID-19 vaccines is often reported by young male adults soon after the second dose; people reporting myocarditis events often recover quickly with no short-term complications.^19^

Hypoaesthesia/paraesthesia and allergy have also been reported as precautions for post-authorization use of NVX-CoV2373.^17^ The frequencies of these events were low and similar for both NVX-CoV2373 and placebo recipients.

## Limitations

One limitation of the study is that the participant populations varied, with residents of the United States, Mexico, South Africa, Australia, and the United Kingdom included. Each of these regions had different COVID-19 precautions in place and were faced with different variants. Additionally, data from post-crossover phases of the trials were excluded due to difficulties in separating the different treatment effects. However, reporting of some AEs may be lost due to exclusion of these data. Further, some participants may have opted to receive an authorized vaccine when they became available; as such, these participants were censored from analyses, which may have introduced selection bias.

To date, NVX-CoV2373 has displayed an acceptable safety profile in healthy adult participants, with the majority of solicited AEs being of mild-to-moderate severity and of short duration. The data shown here support the continued use of NVX-CoV2373 to reduce the risk of COVID-19– associated hospitalization and death.

## Data Availability

The trial protocols and statistical analysis plans will be made available upon publishing of the manuscript by request to the corresponding author. Additional information is available at clinicaltrials.gov.

## Contributors

KS and KH oversaw the analysis of safety data; MRC provided the statistical analysis; IM created and wrote the outline and provided editorial support; MDR provided an epidemiologic analysis; and KA provided a clinical analysis of the data. All authors had access to and verified the underlying data. All authors reviewed and edited multiple drafts and approved the final draft manuscript before submission.

## Acknowledgements

The study and article were funded by Novavax, Inc. We would like to thank all the study participants for their commitment to this study. We also acknowledge the investigators and their study teams for their hard work and dedication. The authors thank Farnaz Mahkou for statistical support in the early phases of the analysis, and Corrie Roncal for her expertise and assistance in pharmacovigilance throughout the 2019nCoV-101 study. Writing and editorial support for this manuscript were funded by Novavax, Inc., and were provided by Rebecca Harris, PhD, and Kelly Cameron, PhD, of Ashfield MedComms (New York, USA), an Inizio company.

