## Supplement for "Safety of the NVX-CoV2373 COVID-19 Vaccine in Randomized Placebo-Controlled Clinical Trials"

Table S1: Description of randomized trials included in this analysis……………………………………………………………......2

Table S2: Frequency of solicited local reactions within 7 days after each dose, by maximum severity and age group …..……………..3

Table S3: Frequency of solicited systemic reactions within 7 days after each dose administration, by maximum severity and age group……………………………………………………………………………………………………………………4

Table S4: Characteristics of solicited local and systemic reactions reported within 7 days after each dose……………………………..5

Table S5: Summary of unsolicited adverse events (occurring in ≥0·1% of participants) reported from Day 0 to 49 (28 days post-Dose 2)…………………………………………………………………………………………………………………..7

Table S6: Subgroup summary of event rates of serious adverse events (≥0·2 events per 100 person-years) reported during the study (from Day 0 to end of follow-up) by age ……………………………………………………………………...….9

Table S7: Summary of adverse events resulting in discontinuation (from Day 0 to crossover) by age……………………….……..11

Table S8: Subgroup summary of event rates of potential immune-mediated medical conditions reported during the study (from Day 0 to end of follow-up) per site or protocol defined criteria, reported by age ……………………………………………….……..14

### Table S1: Description of randomized trials included in this analysis

| Study number (country, registry number) | Study design | Treatment groups (dose 1 at day 0 dose 2 at Day 21) V=SARS CoV-rS vaccine M=Matrix M™ adjuvant P=placebo | Number of participants randomized in study (randomization ratio) | Number of participants included in the analysis | |
|---|---|---|---|---|---|
| | | | | Vaccine (5 µg) | Placebo |
| 2019nCoV-101 Part 1<br><br>(Australia, NCT04368988) | Phase 1, randomized, observer-blinded, placebo-controlled in healthy adults 18–59 years of age | A: P + P<br>B: V (25 µg) and M (0 µg) + V (25 µg) + M (0 µg)<br>C: V (5 µg) and M (50 µg) + V (5 µg) and M (50 µg)<br>D: V (25 µg) and M (50 µg) + V (25 µg) and M (50 µg)<br>E: V (25 µg) and M (50 µg) + P | 134<br>(1:1:1:1:1) | 29<br>(Group C) | 23<br>(Group A) |
| 2019nCoV-101 Part 2<br><br>(Australia and US, NCT04368988) | Phase 2, randomized, observer-blinded, placebo-controlled in healthy adults 18–84 years of age | A: P + P<br>B: V (5 µg) and M (50 µg) + V (5 µg) + M (50 µg)<br>C: V (5 µg) and M (50 µg) + P<br>D: V (25 µg) and M (50 µg) + V (25 µg) and M (50 µg)<br>E: V (25 µg) and M (50 µg) + P | 1288<br>(1:1:1:1:1) | 514 (Groups B and C) | 255 (Group A) |
| 2019nCoV-501<br><br>(South Africa, NCT04533399) | Phase 2a/2b, randomized, observer-blinded, placebo-controlled in healthy adults (HIV-negative) and in medically stable adult HIV-positive adults 18–84 years of age | A: P + P<br>B: V (5 µg) and M (50 µg) + V (5 µg) + M (50 µg) | 4408<br>(1:1) | 2211 | 2197 |
| 2019nCoV-302<br><br>(United Kingdom, EudraCT number, 2020-004123-16) | Phase 3, randomized, observer-blinded, placebo-controlled trial in adults 18–84 years of age | A: P + P<br>B: V (5 µg) and M (50 µg) + V (5 µg) + M (50 µg) | 15,187<br>(1:1) | 7575 | 7564 |
| 2019nCoV-301<br><br>(US and Mexico, NCT04611802) | Phase 3, randomized, observer-blinded, placebo-controlled study in adults ≥18 years of age | A: P + P<br>B: V (5 µg) and M (50 µg) + V (5 µg) + M (50 µg) | 29,949<br>(2:1) | 19,729 | 9853 |
| **Total** | | | **50,964** | **30,058** | **19,892** |

M=Matrix-M™ adjuvant; P=placebo; V=active NVX-CoV2373 vaccine.

**Table S2: Frequency of solicited local reactions within 7 days after each dose, by maximum severity and age group**

| | 18–64 years | | 65–84 years | | All participants | |
|---|---|---|---|---|---|---|
| **Event, n (%)** | **NVX-CoV2373 D1 n=19,434 D2 n=18,272** | **Placebo D1 n=11,128 D2 n=10,454** | **NVX-CoV2373 D1 n=2671 D2 n=2364** | **Placebo D1 n=1497 D2 n=1343** | **NVX-CoV2373 D1 n=22,105 D2 n=20,636** | **Placebo D1 n=12,625 D2 n=11,797** |
| **Any Local AE** | | | | | | |
| Dose 1 (any grade) | 11187 (57·56) | 2294 (20·61) | 991 (37·10) | 216 (14·43) | 12,178 (55·09) | 2510 (19·88) |
| Grade 3+ | 231 (1·19) | 28 (0·25) | 15 (0·56) | 3 (0·20) | 246 (1·11) | 31 (0·25) |
| Dose 2 (any grade) | 13795 (75·50) | 2054 (19·65) | 1418 (59·98) | 187 (13·92) | 15213 (73·72) | 2241 (19·00) |
| Grade 3+ | 1201 (6·57) | 32 (0·31) | 61 (2·58) | 2 (0·15) | 1262 (6·12) | 34 (0·29) |
| **Pain/Tenderness** | | | | | | |
| Dose 1 (any grade) | 11154 (57·39) | 2271 (20·41) | 984 (36·84) | 213 (14·23) | 12138 (54·91) | 2484 (19·68) |
| Grade 3+ | 223 (1·15) | 26 (0·23) | 14 (0·52) | 3 (0·20) | 237 (1·07) | 29 (0·23) |
| Dose 2 (any grade) | 13757 (75·29) | 2033 (19·45) | 1411 (59·69) | 180 (13·40) | 15168 (73·50) | 2213 (18·76) |
| Grade 3+ | 1061 (5·81) | 30 (0·29) | 45 (1·90) | 1 (0·07) | 1106 (5·36) | 31 (0·26) |
| **Erythema** | | | | | | |
| Dose 1 (any grade) | 190 (0·98) | 31 (0·28) | 20 (0·75) | 5 (0·33) | 210 (0·95) | 36 (0·29) |
| Grade 3+ | 4 (0·02) | 0 | 0 | 0 | 4 (0·02) | 0 |
| Dose 2 (any grade) | 1155 (6·32) | 31 (0·30) | 123 (5·20) | 4 (0·30) | 1278 (6·19) | 35 (0·30) |
| Grade 3+ | 145 (0·79) | 2 (0·02) | 8 (0·34) | 0 | 153 (0·74) | 2 (0·02) |
| **Swelling/Induration** | | | | | | |
| Dose 1 (any grade) | 170 (0·87) | 35 (0·31) | 18 (0·67) | 1 (0·07) | 188 (0·85) | 36 (0·29) |
| Grade 3+ | 6 (0·03) | 3 (0·03) | 1 (0·04) | 0 | 7 (0·03) | 3 (0·02) |
| Dose 2 (any grade) | 1061 (5·81) | 26 (0·25) | 136 (5·75) | 7 (0·52) | 1197 (5·80) | 33 (0·28) |
| Grade 3+ | 86 (0·47) | 1 (< 0·01) | 9 (0·38) | 1 (0·07) | 95 (0·46) | 2 (0·02) |

AE=adverse event; D1/D2=Dose 1/Dose 2.

### Table S3: Frequency of solicited systemic reactions within 7 days after each dose administration, by maximum severity and age group

| | 18–64 years | | 65–84 years | | All participants | |
|---|---|---|---|---|---|---|
| **Event** | **NVX-CoV2373 D1 n=19,434 D2 n=18,272** | **Placebo D1 n=11,128 D2 n=10,454** | **NVX-CoV2373 D1 n=2671 D2 n=2364** | **Placebo D1 n=1497 D2 n=1343** | **NVX-CoV2373 D1 n=22,105 D2 n=20,636** | **Placebo D1 n=12,625 D2 n=11,797** |
| **Any Systemic AE** | | | | | | |
| Dose 1 (any grade) | 9233 (47·51) | 4233 (38·04) | 844 (31·60) | 442 (29·53) | 10077 (45·59) | 4675 (37·03) |
| Grade 3+ | 480 (2·47) | 241 (2·17) | 43 (1·61) | 15 (1·00) | 523 (2·37) | 256 (2·03) |
| Dose 2 (any grade) | 12151 (66·50) | 3426 (32·77) | 1101 (46·57) | 341 (25·39) | 13252 (64·22) | 3767 (31·93) |
| Grade 3+ | 2130 (11·66) | 223 (2·13) | 94 (3·98) | 20 (1·49) | 2224 (10·78) | 243 (2·06) |
| **Fever** | | | | | | |
| Dose 1 (Any grade) [≥38·0 °C] | 120 (0·62) | 79 (0·71) | 18 (0·67) | 12 (0·80) | 138 (0·62) | 91 (0·72) |
| Grade 3+ [≥39 °C] | 25 (0·13) | 17 (0·15) | 3 (0·11) | 1 (0·07) | 28 (0·13) | 18 (0·14) |
| Dose 2 (Any grade) [≥38·0 °C] | 1038 (5·68) | 49 (0·47) | 45 (1·90) | 12 (0·89) | 1083 (5·25) | 61 (0·52) |
| Grade 3+ [≥39 °C] | 74 (0·41) | 10 (0·10) | 3 (0·13) | 2 (0·15) | 77 (0·37) | 12 (0·10) |
| **Headache** | | | | | | |
| Dose 1 (any grade) | 4904 (25·23) | 2484 (22·32) | 404 (15·13) | 225 (15·03) | 5308 (24·01) | 2709 (21·46) |
| Grade 3+ | 161 (0·83) | 83 (0·75) | 14 (0·52) | 4 (0·27) | 175 (0·79) | 87 (0·69) |
| Dose 2 (any grade) | 7902 (43·25) | 1932 (18·48) | 553 (23·39) | 173 (12·88) | 8455 (40·97) | 2105 (17·84) |
| Grade 3+ | 556 (3·04) | 67 (0·64) | 19 (0·80) | 3 (0·22) | 575 (2·79) | 70 (0·59) |
| **Fatigue/Malaise** | | | | | | |
| Dose 1 (any grade) | 5591 (28·77) | 2615 (23·50) | 501 (18·76) | 244 (16·30) | 6092 (27·56) | 2859 (22·65) |
| Grade 3+ | 294 (1·51) | 135 (1·21) | 26 (0·97) | 7 (0·47) | 320 (1·45) | 142 (1·12) |
| Dose 2 (any grade) | 9633 (52·72) | 2261 (21·63) | 769 (32·53) | 218 (16·23) | 10402 (50·41) | 2479 (21·01) |
| Grade 3+ | 1668 (9·13) | 143 (1·37) | 68 (2·88) | 17 (1·27) | 1736 (8·41) | 160 (1·36) |
| **Myalgia (Muscle Pain)** | | | | | | |
| Dose 1 (any grade) | 4423 (22·76) | 1417 (12·73) | 333 (12·47) | 150 (10·02) | 4756 (21·52) | 1567 (12·41) |
| Grade 3+ | 104 (0·54) | 43 (0·39) | 3 (0·11) | 4 (0·27) | 107 (0·48) | 47 (0·37) |
| Dose 2 (any grade) | 8397 (45·96) | 1116 (10·68) | 615 (26·02) | 123 (9·16) | 9012 (43·67) | 1239 (10·50) |
| Grade 3+ | 869 (4·76) | 47 (0·45) | 33 (1·40) | 3 (0·22) | 902 (4·37) | 50 (0·42) |
| **Arthralgia (Joint Pain)** | | | | | | |
| Dose 1 (any grade) | 1545 (7·95) | 735 (6·60) | 164 (6·14) | 90 (6·01) | 1709 (7·73) | 825 (6·53) |
| Grade 3+ | 67 (0·34) | 30 (0·27) | 6 (0·22) | 5 (0·33) | 73 (0·33) | 35 (0·28) |
| Dose 2 (any grade) | 3910 (21·40) | 667 (6·38) | 292 (12·35) | 76 (5·66) | 4202 (20·36) | 743 (6·30) |
| Grade 3+ | 441 (2·41) | 33 (0·32) | 20 (0·85) | 3 (0·22) | 461 (2·23) | 36 (0·31) |
| **Nausea/Vomiting** | | | | | | |
| Dose 1 (any grade) | 1282 (6·60) | 639 (5·74) | 99 (3·71) | 37 (2·47) | 1381 (6·25) | 676 (5·35) |
| Grade 3+ | 28 (0·14) | 17 (0·15) | 0 | 0 | 28 (0·13) | 17 (0·13) |
| Dose 2 (any grade) | 2059 (11·27) | 545 (5·21) | 121 (5·12) | 41 (3·05) | 2180 (10·56) | 586 (4·97) |
| Grade 3+ | 46 (0·25) | 16 (0·15) | 2 (0·08) | 0 | 48 (0·23) | 16 (0·14) |

AE=adverse event; D1/D2=Dose 1/Dose 2.

**Table S4: Characteristics of solicited local and systemic reactions reported within 7 days after each dose**

| | Dose 1 | | Dose 2 | |
|---|---|---|---|---|
| **Event*** | **NVX-CoV2373 N=22,105** | **Placebo N=12,625** | **NVX-CoV2373 N =20,636** | **Placebo N=11,797** |
| Any local reaction | | | | |
| Day of onset, median (min – max) | 2 (1–7) | 1 (1–7) | 2 (1–7) | 1 (1–7) |
| Duration, median (min – max) | 2 (1–7) | 1 (1–7) | 2 (1–7) | 1 (1–7) |
| Persisted beyond Day 7, n | 18 | 4 | 72 | 2 |
| Erythema | | | | |
| Day of onset, median (min – max) | 2 (1–7) | 2 (1–7) | 3 (1–7) | 2 (1–7) |
| Duration, median (min – max) | 1 (1–7) | 1 (1–5) | 2 (1–7) | 1 (1–6) |
| Persisted beyond Day 7, n | 2 | 1 | 19 | 1 |
| Pain/Tenderness | | | | |
| Day of onset, median (min – max) | 2 (1–7) | 1 (1–7) | 2 (1–7) | 1 (1–7) |
| Duration, median (min – max) | 2 (1–7) | 1 (1–7) | 3 (1–7) | 1 (1–7) |
| Persisted beyond Day 7, n | 12 | 2 | 36 | 1 |
| Swelling/Induration | | | | |
| Day of onset, median (min – max) | 2 (1–7) | 1·5 (1,7) | 2 (1–7) | 2 (1–7) |
| Duration, median (min – max) | 1 (1–7) | 1 (1–5) | 2 (1–7) | 1 (1–5) |
| Persisted beyond Day 7, n | 4 | 1 | 17 | 0 |
| Any systemic reaction | | | | |
| Day of onset, median (min – max) | 2 (1–7) | 2 (1–7) | 2 (1–7) | 2 (1–7) |
| Duration, median (min – max) | 1 (1–7) | 1 (1–7) | 1 (1–7) | 1 (1–7) |
| Persisted beyond Day 7, n | 85 | 47 | 130 | 65 |
| Fatigue/Malaise | | | | |
| Day of onset, median (min – max) | 2 (1–7) | 2 (1–7) | 2 (1–7) | 2 (1–7) |
| Duration, median (min – max) | 1 (1–7) | 1 (1–7) | 1 (1–7) | 1 (1–7) |
| Persisted beyond Day 7, n | 22 | 5 | 22 | 18 |
| Fever | | | | |
| Day of onset, median (min – max) | 4 (1–7) | 4 (1–7) | 2 (1–7) | 4 (1–7) |
| Duration, median (min – max) | 1 (1–6) | 1 (1–4) | 1 (1–6) | 1 (1–4) |
| Persisted beyond Day 7, n | 3 | 1 | 7 | 0 |
| Headache | | | | |
| Day of onset, median (min – max) | 2 (1–7) | 2 (1–7) | 2 (1–7) | 2 (1–7) |
| Duration, median (min – max) | 1 (1–7) | 1 (1–7) | 1 (1–7) | 1 (1–7) |
| Persisted beyond Day 7, n | 32 | 21 | 52 | 26 |
| Arthralgia (Joint Pain) | | | | |
| Day of onset, median (min – max) | 2 (1–7) | 3 (1–7) | 2 (1–7) | 2 (1–7) |
| Duration, median (min – max) | 1 (1–7) | 1 (1–7) | 1 (1–7) | 1 (1–7) |
| Persisted beyond Day 7, n | 7 | 8 | 16 | 6 |
| Myalgia (Muscle Pain) | | | | |
| Day of onset, median (min – max) | 2 (1–7) | 2 (1–7) | 2 (1–7) | 2 (1–7) |
| Duration, median (min – max) | 1 (1–7) | 1 (1–7) | 1 (1–7) | 1 (1–7) |
| Persisted beyond Day 7, n | 13 | 11 | 20 | 11 |
| Nausea/Vomiting | | | | |
| Day of onset, median (min – max) | 3 (1–7) | 3 (1–7) | 2 (1–7) | 2 (1–7) |
| Duration, median (min – max) | 1 (1–7) | 1 (1–7) | 1 (1–7) | 1 (1–7) |

| | | | | |
|---|---|---|---|---|
| Persisted beyond Day 7, n | 8 | 1 | 13 | 4 |

Max=maximum; min=minimum.
*Participants were followed for solicited events for 7 days following each dose administration.

**Table S5: Summary of unsolicited adverse events (occurring in ≥0·1% of participants) reported from Day 0 to 49 (28 days post-Dose 2)**

| System organ class<br>preferred term (# of participants) | NVX-CoV2373<br>n (%)<br>N=30,072 | Placebo<br>n (%)<br>N=19,877 | Risk difference<br>(vaccine – placebo)<br>% (95% CI) |
|---|---|---|---|
| **Any unsolicited AEs** | 6110 (20·32) | 3448 (17·35) | 5·85 (5·16, 6·54) |
| **General disorders and administration site conditions** | 2168 (7·21) | 670 (3·37) | 5·50 (5·10, 5·90) |
| Injection site pain | 681 (2·26) | 99 (0·50) | 2·38 (2·16, 2·61) |
| Fatigue | 572 (1·90) | 282 (1·42) | 1·00 (0·77, 1·24) |
| Pyrexia | 279 (0·93) | 59 (0·30) | 0·82 (0·67, 0·97) |
| Chills | 178 (0·59) | 23 (0·12) | 0·56 (0·45, 0·66) |
| Pain | 146 (0·49) | 49 (0·25) | 0·32 (0·21, 0·44) |
| Injection site erythema | 132 (0·44) | 15 (0·08) | 0·45 (0·36, 0·55) |
| Injection site swelling | 119 (0·40) | 8 (0·04) | 0·44 (0·35, 0·53) |
| Malaise | 120 (0·40) | 39 (0·20) | 0·31 (0·21, 0·41) |
| Injection site pruritus | 92 (0·31) | 7 (0·04) | 0·32 (0·25, 0·40) |
| Influenza like illness | 89 (0·30) | 18 (0·09) | 0·28 (0·19, 0·36) |
| Vaccination site pain | 42 (0·14) | 13 (0·07) | 0·12 (0·06, 0·18) |
| Injection site bruising | 38 (0·13) | 15 (0·08) | 0·09 (0·03, 0·15) |
| **Nervous system disorders** | 1355 (4·51) | 812 (4·09) | 1·39 (1·02, 1·76) |
| Headache | 947 (3·15) | 529 (2·66) | 1·24 (0·93, 1·55) |
| Lethargy | 109 (0·36) | 40 (0·20) | 0·29 (0·19, 0·39) |
| Dizziness | 105 (0·35) | 66 (0·33) | 0·07 (-0·04, 0·18) |
| Migraine | 41 (0·14) | 23 (0·12) | 0·05 (-0·02, 0·11) |
| Paraesthesia | 31 (0·10) | 28 (0·14) | -0·02 (-0·09, 0·04) |
| Ageusia | 22 (0·07) | 32 (0·16) | -0·06 (-0·13, -0·00) |
| Anosmia | 20 (0·07) | 37 (0·19) | -0·10 (-0·16, -0·03) |
| **Musculoskeletal and connective tissue disorders** | 1313 (4·37) | 497 (2·50) | 2·82 (2·48, 3·16) |
| Myalgia | 498 (1·66) | 140 (0·70) | 1·39 (1·18, 1·59) |
| Pain in extremity | 391 (1·30) | 67 (0·34) | 1·33 (1·15, 1·50) |
| Arthralgia | 194 (0·65) | 110 (0·55) | 0·22 (0·08, 0·36) |
| Back pain | 92 (0·31) | 72 (0·36) | -0·00 (-0·11, 0·10) |
| Musculoskeletal stiffness | 33 (0·11) | 11 (0·06) | 0·08 (0·03, 0·14) |
| Neck pain | 26 (0·09) | 25 (0·13) | -0·02 (-0·08, 0·04) |
| **Infections and infestations** | 891 (2·96) | 677 (3·41) | -0·30 (-0·63, 0·02) |
| Upper respiratory tract infection | 125 (0·42) | 66 (0·33) | 0·10 (-0·02, 0·21) |
| Urinary tract infection | 91 (0·30) | 64 (0·32) | 0·01 (-0·09, 0·11) |
| COVID-19 | 56 (0·19) | 59 (0·30) | -0·16 (-0·26, -0·07) |
| Sinusitis | 44 (0·15) | 40 (0·20) | -0·06 (-0·14, 0·02) |
| Viral infection | 43 (0·14) | 23 (0·12) | -0·00 (-0·07, 0·06) |
| Gastroenteritis | 30 (0·10) | 27 (0·14) | -0·02 (-0·08, 0·05) |
| Nasopharyngitis | 29 (0·10) | 29 (0·15) | -0·02 (-0·08, 0·05) |
| Tooth abscess | 21 (0·07) | 23 (0·12) | -0·03 (-0·09, 0·03) |
| Tooth infection | 20 (0·07) | 32 (0·16) | -0·08 (-0·15, -0·02) |
| Oral herpes | 16 (0·05) | 21 (0·11) | -0·03 (-0·08, 0·02) |
| **Gastrointestinal disorders** | 674 (2·24) | 443 (2·23) | 0·29 (0·02, 0·56) |

| | | | |
|---|---|---|---|
| Diarrhoea | 202 (0·67) | 143 (0·72) | 0·01 (-0·14, 0·16) |
| Nausea | 198 (0·66) | 124 (0·62) | 0·14 (-0·00, 0·29) |
| Vomiting | 61 (0·20) | 34 (0·17) | 0·06 (-0·02, 0·14) |
| Gastrooesophageal reflux disease | 38 (0·13) | 23 (0·12) | 0·03 (-0·04, 0·09) |
| Abdominal pain | 37 (0·12) | 21 (0·11) | 0·04 (-0·02, 0·10) |
| Abdominal pain upper | 30 (0·10) | 15 (0·08) | 0·03 (-0·03, 0·08) |
| Toothache | 26 (0·09) | 22 (0·11) | -0·01 (-0·07, 0·05) |
| **Respiratory, thoracic and mediastinal disorders** | 669 (2·22) | 497 (2·50) | -0·04 (-0·32, 0·23) |
| Nasal congestion | 177 (0·59) | 112 (0·56) | -0·03 (-0·16, 0·11) |
| Oropharyngeal pain | 174 (0·58) | 139 (0·70) | 0·02 (-0·13, 0·16) |
| Cough | 150 (0·50) | 116 (0·58) | -0·05 (-0·18, 0·08) |
| Rhinorrhoea | 121 (0·40) | 110 (0·55) | -0·08 (-0·20, 0·04) |
| Dyspnoea | 57 (0·19) | 38 (0·19) | 0·01 (-0·07, 0·09) |
| Asthma | 19 (0·06) | 19 (0·10) | -0·02 (-0·07, 0·03) |
| **Skin and subcutaneous tissue disorders** | 402 (1·34) | 202 (1·02) | 0·46 (0·27, 0·66) |
| Rash | 81 (0·27) | 42 (0·21) | 0·06 (-0·03, 0·15) |
| Pruritus | 56 (0·19) | 19 (0·10) | 0·12 (0·05, 0·19) |
| Erythema | 31 (0·10) | 5 (0·03) | 0·09 (0·04, 0·14) |
| **Injury, poisoning and procedural complications** | 326 (1·08) | 218 (1·10) | 0·02 (-0·17, 0·21) |
| Contusion | 29 (0·10) | 15 (0·08) | 0·03 (-0·02, 0·08) |
| Ligament sprain | 18 (0·06) | 20 (0·10) | -0·04 (-0·10, 0·01) |
| **Vascular disorders** | 222 (0·74) | 131 (0·66) | 0·15 (-0·00, 0·30) |
| Hypertension | 158 (0·53) | 112 (0·56) | 0·02 (-0·11, 0·16) |
| **Psychiatric disorders** | 179 (0·60) | 109 (0·55) | 0·07 (-0·07, 0·20) |
| Anxiety | 54 (0·18) | 32 (0·16) | 0·02 (-0·05, 0·10) |
| Depression | 37 (0·12) | 25 (0·13) | 0·00 (-0·06, 0·07) |
| Insomnia | 30 (0·10) | 23 (0·12) | -0·01 (-0·07, 0·05) |
| **Blood and lymphatic system disorders** | 168 (0·56) | 77 (0·39) | 0·24 (0·12, 0·37) |
| Lymphadenopathy | 129 (0·43) | 57 (0·29) | 0·22 (0·11, 0·32) |
| **Investigations** | 146 (0·49) | 92 (0·46) | 0·10 (-0·03, 0·22) |
| Blood pressure increased | 43 (0·14) | 29 (0·15) | 0·03 (-0·04, 0·10) |
| **Metabolism and nutrition disorders** | 133 (0·44) | 82 (0·41) | 0·03 (-0·09, 0·15) |
| **Eye disorders** | 100 (0·33) | 59 (0·30) | 0·08 (-0·02, 0·18) |
| **Ear and labyrinth disorders** | 92 (0·31) | 50 (0·25) | 0·09 (-0·01, 0·18) |
| **Reproductive system and breast disorders** | 90 (0·30) | 52 (0·26) | 0·05 (-0·05, 0·14) |
| **Cardiac disorders** | 77 (0·26) | 50 (0·25) | 0·02 (-0·07, 0·11) |
| **Renal and urinary disorders** | 43 (0·14) | 26 (0·13) | 0·01 (-0·06, 0·08) |
| **Immune system disorders** | 38 (0·13) | 16 (0·08) | 0·04 (-0·01, 0·10) |
| **Neoplasms benign, malignant and unspecified (incl cysts and polyps)** | 34 (0·11) | 27 (0·14) | -0·01 (-0·07, 0·05) |

AE=adverse event.

**Table S6: Subgroup summary of event rates of serious adverse events (≥0·2 events per 100 person-years) reported during the study (from Day 0 to end of follow-up) by age**

| | Participants 18–64 years | | Participants ≥65 years | | All participants | |
|---|---|---|---|---|---|---|
| **Event (system organ class preferred term [# of events])** | **NVX-CoV2373 (event n/subject n, rate per 100 person-years) N=25300** | **Placebo (event n/subject n, rate per 100 person-years) N=16413** | **NVX-CoV2373 (event n/subject n, rate per 100 person-years) N=4777** | **Placebo (event n/subject n, rate per 100 person-years) N=3458** | **NVX-CoV2373 (event n/subject n, rate per 100 person-years) N =30,077** | **Placebo (event n/subject n, rate per 100 person-years) N=19,871** |
| **Any SAEs** | 260/205 (2·612) | 185/149 (2·719) | 92/70 (5·307) | 64/49 (5·261) | 350/275 (7·919) | 249/198 (7·98) |
| **Infections and infestations** | 48/43 (0·548) | 51/48 (0·876) | 12/10 (0·758) | 16/12 (1·288) | 60/53 (0·58) | 67/60 (0·94) |
| COVID-19 | 5/5 (0·064) | 6/6 (0·109) | 2/2 (0·152) | 3/3 (0·322) | 7/7 (0·08) | 9/9 (0·14) |
| Pneumonia | 2/2 (0·025) | 4/4 (0·073) | 3/3 (0·227) | 4/3 (0·322) | 5/5 (0·05) | 8/7 (0·11) |
| COVID-19 pneumonia | 2/2 (0·025) | 13/13 (0·237) | 0 | 2/2 (0·215) | 2/2 (0·02) | 15/15 (0·23) |
| **Cardiac disorders** | 25/23 (0·293) | 13/13 (0·237) | 16/15 (1·137) | 8/8 (0·859) | 41/38 (0·41) | 21/21 (0·33) |
| Acute myocardial infarction | 3/3 (0·038) | 2/2 (0·036) | 2/2 (0·152) | 1/1 (0·107) | 5/5 (0·05) | 3/3 (0·05) |
| Myocardial infarction | 1/1 (0·013) | 1/1 (0·018) | 1/1 (0·076) | 2/2 (0·215) | 2/2 (0·02) | 3/3 (0·05) |
| **Injury, poisoning and procedural complications** | 35/30 (0·382) | 23/21 (0·383) | 14/13 (0·986) | 6/6 (0·644) | 49/43 (0·47) | 29/27 (0·42) |
| **Renal and urinary disorders** | 4/4 (0·051) | 5/5 (0·091) | 6/6 (0·455) | 2/2 (0·215) | 10/10 (0·11) | 7/7 (0·11) |
| Acute kidney injury | 3/3 (0·038) | 1/1 (0·018) | 5/5 (0·379) | 2/2 (0·215) | 8/8 (0·09) | 3/3 (0·05) |
| **Nervous system disorders** | 23/21 (0·268) | 15/15 (0·274) | 5/5 (0·379) | 1/1 (0·107) | 28/26 (0·28) | 16/16 (0·25) |
| Cerebrovascular accident | 4/4 (0·051) | 2/2 (0·036) | 3/3 (0·227) | 1/1 (0·107) | 7/7 (0·08) | 3/3 (0·05) |
| **Gastrointestinal disorders** | 24/20 (0·255) | 4/4 (0·073) | 3/3 (0·227) | 3/3 (0·322) | 27/23 (0·25) | 7/7 (0·11) |
| **Psychiatric disorders** | 15/14 (0·178) | 12/11 (0·201) | 0 | 4/3 (0·322) | 15/14 (0·15) | 16/14 (0·22) |
| **Respiratory, thoracic and mediastinal disorders** | 12/10 (0·127) | 6/6 (0·109) | 7/6 (0·455) | 5/5 (0·537) | 19/16 (0·17) | 11/11 (0·17) |
| Pulmonary embolism | 2/2 (0·025) | 3/3 (0·055) | 2/2 (0·152) | 2/2 (0·215) | 4/4 (0·04) | 5/5 (0·08) |
| **Neoplasms benign, malignant and unspecified (incl cysts and polyps)** | 16/16 (0·204) | 9/9 (0·164) | 12/12 (0·910) | 9/9 (0·966) | 28/28 (0·31) | 18/18 (0·28) |
| Prostate cancer | 2/2 (0·025) | 0 | 5/5 (0·379) | 0 | 7/7 (0·08) | 0 |
| **General disorders and administration site conditions** | 3/3 (0·038) | 3/3 (0·055) | 5/5 (0·379) | 3/3 (0·322) | 8/8 (0·09) | 6/6 (0·09) |
| Death | 0 | 0 | 3/3 (0·227) | 0 | 3/3 (0·03) | 0 |

| | | | | | | |
|---|---|---|---|---|---|---|
| **Musculoskeletal and connective tissue disorders** | 5/5 (0·064) | 4/4 (0·073) | 3/3 (0·227) | 0 | 8/8 (0·09) | 4/4 (0·06) |
| **Vascular disorders** | 8/8 (0·102) | 4/4 (0·073) | 3/2 (0·152) | 3/3 (0·322) | 11/10 (0·11) | 7/7 (0·11) |

SAE=serious adverse event.

**Table S7: Summary of adverse events resulting in discontinuation (from Day 0 to crossover) by age**

| | Participants 18–64 years | | Participants ≥65 years | |
|---|---|---|---|---|
| **System organ class<br>preferred term [# of events]** | **NVX-CoV2373<br>(event n/subject n, rate per 100 person-years)<br>N=25300** | **Placebo<br>(event n/subject n, rate per 100 person-years)<br>N=16413** | **NVX-CoV2373<br>(event n/subject n, rate per 100 person-years)<br>N=4777** | **Placebo<br>(event n/subject n, rate per 100 person-years)<br>N=3458** |
| **Number of participants experiencing any event** | 65/61 (0·777) | 24/23 (0·420) | 24/21 (1·592) | 13/13 (1·396) |
| **Cardiac disorders** | 6/6 (0·076) | 5/5 (0·091) | 4/4 (0·303) | 4/4 (0·429) |
| Acute myocardial infarction | 1/1 (0·013) | 0 | 0 | 0 |
| Angina pectoris | 0 | 0 | 0 | 1/1 (0·107) |
| Arrythmia | 0 | 0 | 0 | 1/1 (0·107) |
| Atrial fibrillation | 1/1 (0·013) | 0 | 0 | 1/1 (0·107) |
| Atrial flutter | 0 | 1/1 (0·018) | 0 | 0 |
| Bradycardia | 0 | 1/1 (0·018) | 1/1 (0·076) | 0 |
| Cardiac arrest | 4/4 (0·051) | 2/2 (0·036) | 1/1 (0·076) | 0 |
| Cardio-respiratory arrest | 0 | 1/1 (0·018) | 0 | 0 |
| Myocardial infarction | 0 | 0 | 0 | 1/1 (0·107) |
| Sinus bradycardia | 0 | 0 | 1/1 (0·076) | 0 |
| Tachycardia | 0 | 0 | 1/1 (0·076) | 0 |
| **Endocrine disorders** | 2/2 (0·025) | 0 | 0 | 0 |
| Autoimmune thyroiditis | 1/1 (0·013) | 0 | 0 | 0 |
| Hypothyroidism | 1/1 (0·013) | 0 | 0 | 0 |
| **Eye disorders** | 0 | 1/1 (0·018) | 0 | 0 |
| Conjunctivitis allergic | 0 | 1/1 (0·018) | 0 | 0 |
| **Gastrointestinal disorders** | 2/2 (0·025) | 1/1 (0·018) | 1/1 (0·076) | 1/1 (0·107) |
| Abdominal pain | 1/1 (0·013) | 0 | 0 | 0 |
| Colitis | 0 | 1/1 (0·018) | 0 | 0 |
| Diarrhea | 1/1 (0·013) | 0 | 1/1 (0·076) | 0 |
| Nausea | 0 | 0 | 0 | 1/1 (0·107) |
| **General disorders and administration site conditions** | 10/10 (0·127) | 1/1 (0·018) | 6/6 (0·455) | 1/1 (0·107) |
| Asthenia | 0 | 0 | 1/1 (0·076) | 0 |
| Chest discomfort | 1/1 (0·013) | 0 | 0 | 0 |
| Chills | 0 | 0 | 1/1 (0·076) | 0 |
| Death | 0 | 0 | 3/3 (0·227) | 0 |
| Fatigue | 2/2 (0·025) | 0 | 0 | 0 |
| Influenza like illness | 0 | 0 | 0 | 1/1 (0·107) |
| Injection site erythema | 1/1 (0·013) | 0 | 0 | 0 |
| Injection site pain | 5/5 (0·064) | 0 | 1/1 (0·076) | 0 |
| Injection site pruritis | 1/1 (0·013) | 0 | 0 | 0 |
| Systemic inflammatory response syndrome | 0 | 1/1 (0·018) | 0 | 0 |
| **Hepatobiliary disorders** | 1/1 (0·013) | 0 | 0 | 0 |
| Bile duct stone | 1/1 (0·013) | 0 | 0 | 0 |
| **Infections and Infestations** | 10/10 (0·127) | 4/4 (0·073) | 1/1 (0·076) | 3/3 (0·322) |
| Appendicitis perforated | 1/1 (0·013) | 0 | 0 | 0 |
| COVID-19 | 1/1 (0·013) | 2/2 (0·036) | 1/1 (0·076) | 1/1 (0·107) |
| COVID-19 pneumonia | 1/1 (0·013) | 1/1 (0·018) | 0 | 0 |
| Empyema | 1/1 (0·013) | 0 | 0 | 0 |

| | | | | |
|---|---|---|---|---|
| Enterococcal sepsis | 0 | 0 | 0 | 1/1 (0·107) |
| Hepatitis B | 1/1 (0·013) | 0 | 0 | 0 |
| Infectious mononucleosis | 1/1 (0·013) | 0 | 0 | 0 |
| Pharyngitis streptococcal | 1/1 (0·013) | 0 | 0 | 0 |
| Respiratory tract infection | 0 | 1/1 (0·018) | 0 | 0 |
| Septic shock | 1/1 (0·013) | 0 | 0 | 0 |
| Suspected COVID-19 | 1/1 (0·013) | 0 | 0 | 0 |
| Vulvitis | 1/1 (0·013) | 0 | 0 | 0 |
| Vulvovaginal candidiasis | 0 | 0 | 0 | 1/1 (0·107) |
| **Injury, poisoning and procedural complications** | 6/6 (0·076) | 2/2 (0·036) | 3/3 (0·227) | 0 |
| Accidental overdose | 1/1 (0·013) | 0 | 0 | 0 |
| Alcohol poisoning | 1/1 (0·013) | 0 | 0 | 0 |
| Fall | 0 | 0 | 2/2 (0·152) | 0 |
| Gunshot wound | 2/2 (0·025) | 0 | 0 | 0 |
| Intentional overdose | 1/1 (0·013) | 0 | 0 | 0 |
| Multiple injuries | 0 | 1/1 (0·018) | 0 | 0 |
| Overdose | 0 | 1/1 (0·018) | 0 | 0 |
| Poisoning deliberate | 0 | 0 | 1/1 (0·076) | 0 |
| Toxicity to various agents | 1/1 (0·013) | 0 | 0 | 0 |
| **Investigations** | 1/1 (0·013) | 0 | 3/3 (0·227) | 0 |
| Antinuclear antibody positive | 1/1 (0·013) | 0 | 0 | 0 |
| Blood pressure increased | 0 | 0 | 1/1 (0·076) | 0 |
| Cardiac murmur | 0 | 0 | 1/1 (0·076) | 0 |
| Heart rate increased | 0 | 0 | 1/1 (0·076) | 0 |
| **Metabolism and nutrition disorders** | 2/2 (0·025) | 0 | 0 | 0 |
| Diabetic ketoacidosis | 1/1 (0·013) | 0 | 0 | 0 |
| Type 2 diabetes mellitus | 1/1 (0·013) | 0 | 0 | 0 |
| **Musculoskeletal and connective tissue disorders** | 5/5 (0·064) | 0 | 1/1 (0·076) | 0 |
| Myalgia | 2/2 (0·025) | 0 | 1/1 (0·076) | 0 |
| Neck pain | 1/1 (0·013) | 0 | 0 | 0 |
| Pain in extremity | 1/1 (0·013) | 0 | 0 | 0 |
| Rheumatoid arthritis | 1/1 (0·013) | 0 | 0 | 0 |
| **Neoplasms benign, malignant and unspecified (incl cysts and polyps)** | 0 | 0 | 1/1 (0·076) | 3/3 (0·322) |
| Glioblastoma | 0 | 0 | 0 | 1/1 (0·107) |
| Metastases to liver | 0 | 0 | 1/1 (0·076) | 0 |
| Non-Hodgkin's lymphoma | 0 | 0 | 0 | 1/1 (0·107) |
| Ovarian cancer | 0 | 0 | 0 | 1/1 (0·107) |
| **Nervous system disorders** | 6/6 (0·076) | 3/3 (0·055) | 2/2 (0·152) | 0 |
| Cerebrovascular accident | 1/1 (0·013) | 0 | 1/1 (0·076) | 0 |
| Dizziness | 0 | 0 | 1/1 (0·076) | 0 |
| Headache | 1/1 (0·013) | 0 | 0 | 0 |
| Hypoaesthesia | 1/1 (0·013) | 0 | 0 | 0 |
| Migraine with aura | 0 | 1/1 (0·018) | 0 | 0 |
| Multiple sclerosis | 0 | 1/1 (0·018) | 0 | 0 |
| Paraesthesia | 2/2 (0·025) | 0 | 0 | 0 |
| Seizure | 0 | 1/1 (0·018) | 0 | 0 |
| Somnolence | 1/1 (0·013) | 0 | 0 | 0 |

| | | | | |
|---|---|---|---|---|
| **Pregnancy, puerperium and perinatal conditions** | 2/2 (0·025) | 0 | 0 | 0 |
| Abortion spontaneous | 1/1 (0·013) | 0 | 0 | 0 |
| Pregnancy | 1/1 (0·013) | 0 | 0 | 0 |
| **Psychiatric disorders** | 4/3 (0·038) | 2/2 (0·036) | 0 | 0 |
| Anxiety | 0 | 1/1 (0·018) | 0 | 0 |
| Completed suicide | 1/1 (0·013) | 1/1 (0·018) | 0 | 0 |
| Drug abuse | 1/1 (0·013) | 0 | 0 | 0 |
| Homicidal ideation | 1/1 (0·013) | 0 | 0 | 0 |
| Substance-induced psychotic disorder | 1/1 (0·013) | 0 | 0 | 0 |
| **Renal and urinary disorders** | 0 | 0 | 1/1 (0·076) | 0 |
| Urinary incontinence | 0 | 0 | 1/1 (0·076) | 0 |
| **Respiratory, thoracic and mediastinal disorders** | 3/3 (0·038) | 3/2 (0·036) | 0 | 1/1 (0·107) |
| Asthma | 0 | 0 | 0 | 1/1 (0·107) |
| Cough | 1/1 (0·013) | 2/2 (0·036) | 0 | 0 |
| Nasal congestion | 0 | 1/1 (0·018) | 0 | 0 |
| Pharyngeal hypoaesthesia | 1/1 (0·013) | 0 | 0 | 0 |
| Pulmonary embolism | 1/1 (0·013) | 0 | 0 | 0 |
| **Skin and subcutaneous tissue disorders** | 2/1 (0·013) | 0 | 0 | 0 |
| Angioedema | 1/1 (0·013) | 0 | 0 | 0 |
| Urticaria | 1/1 (0·013) | 0 | 0 | 0 |
| **Vascular disorders** | 3/3 (0·038) | 2/2 (0·036) | 1/1 (0·076) | 0 |
| Hypertension | 3/3 (0·038) | 2/2 (0·036) | 1/1 (0·076) | 0 |

**Table S8. Subgroup summary of event rates of potential immune-mediated medical conditions reported during the study (from Day 0 to end of follow-up) per site or protocol defined criteria, reported by age**

| | Age 18–64 years | | | Age ≥65 years | | | All participants | | |
|---|---|---|---|---|---|---|---|---|---|
| **System organ class preferred term (# of events)** | **NVX-CoV2373 (n, rate per 100 person-years, 95% CI) N=25282** | **Placebo (n, rate per 100 person-years, 95% CI) N=16433** | **Risk difference* (vaccine - placebo) (rate per 100 person-years, 95% CI)** | **NVX-CoV2373 (n, rate per 100 person-years, 95% CI) N =4776** | **Placebo (n, rate per 100 person-years, 95% CI) N=3459** | **Risk difference (vaccine - placebo) (rate per 100 person-years, 95% CI)** | **NVX-CoV2373 (n, rate per 100 person-years, 95% CI) N=30,058** | **Placebo (n, rate per 100 person-years, 95% CI) N=19,892** | **Risk difference (vaccine - placebo) (rate per 100 person-years, 95% CI)** |
| **Total follow-up time (person-years)** | 6337·9 | 4074·4 | - | 1127·1 | 802·8 | - | 7465.0 | 4877·1 | - |
| **Average follow-up time (days)** | 91·6 | 90·6 | - | 86·2 | 84·8 | - | 90·7 | 89·6 | - |
| **Median follow-up time (days)** | 93 | 92 | - | 91 | 88 | - | 92 | 91 | - |
| **Any PIMMCs** | 36 (0·57), (0·40, 0·79) | 16 (0·39), (0·22, 0·64) | 0·11 (-0·17, 0·39) | 5 (0·44), (0·14, 1·04) | 5 (0·62), (0·20, 1·45) | -0·24 (-0·90, 0·42) | 41 (0·55), (0·39, 0·75) | 21 (0·43), (0·27, 0·66) | 0·06 (-0·20, 0·31) |
| **Nervous system disorders** | 12 (0·19), (0·10, 0·33) | 6 (0·15), (0·05, 0·32) | 0·01 (-0·16, 0·17) | 1 (0·09), (0·00, 0·49) | 2 (0·25), (0·03, 0·90) | -0·23 (-0·67, 0·21) | 13 (0·17), (0·09, 0·30) | 8 (0·16), (0·07, 0·32) | -0·03 (-0·18, 0·12) |
| Seizure | 4 (0·06) | 3 (0·07) | -0·03 (-0·14, 0·08) | 0 | 0 | 0 | 4 (0·05) | 3 (0·06) | -0·03 (-0·12, 0·07) |
| Neuropathy peripheral | 3 (0·05) | 0 | 0·04 (-0·01, 0·09) | 0 | 2 (0·25) | -0·30 (-0·72, 0·12) | 3 (0·04) | 2 (0·04) | -0·01 (-0·09, 0·06) |
| Central nervous system inflammation | 1 (0·02) | 0 | 0·01 (-0·01, 0·04) | 0 | 0 | 0 | 1 (0·01) | 0 | 0·01 (-0·01, 0·03) |
| Facial paralysis | 1 (0·02) | 1 (0·02) | 0 (-0·06, 0·06) | 0 | 0 | 0 | 1 (0·01) | 1 (0·02) | 0 (-0·05, 0·05) |
| Hypoaesthesia | 1 (0·02) | 1 (0·02) | -0·01 (-0·08, 0·05) | 0 | 0 | 0 | 1 (0·01) | 1 (0·02) | -0·01 (-0·06, 0·04) |
| Narcolepsy | 1 (0·02) | 0 | 0·01 (-0·01, 0·04) | 0 | 0 | 0 | 1 (0·01) | 0 | 0·01 (-0·01, 0·03) |
| Neuralgia | 0 | 0 | 0 | 1 (0·09) | 0 | 0·07 (-0·07, 0·21) | 1 (0·01) | 0 | 0·01 (-0·01, 0·03) |
| Peroneal nerve palsy | 1 (0·02) | 0 | 0·01 (-0·01, 0·04) | 0 | 0 | 0 | 1 (0·01) | 0 | 0·01 (-0·01, 0·03) |
| Multiple sclerosis | 0 | 1 (0·02) | -0·03 (-0·08, 0·03) | 0 | 0 | 0 | 0 | 1 (0·02) | -0·02 (-0·07, 0·02) |
| **Musculoskeletal and connective tissue disorders** | 5 (0·08), (0·03, 0·18) | 3 (0·07), (0·02, 0·22) | 0·01 (-0·11, 0·13) | 2 (0·18), (0·02, 0·64) | 2 (0·25), (0·03, 0·90) | -0·04 (-0·44, 0·36) | 7 (0·09), (0·04, 0·19) | 5 (0·10), (0·03, 0·24) | 0 (-0·11, 0·12) |
| Arthritis | 2 (0·03) | 0 | 0·03 (-0·01, 0·08) | 0 | 0 | 0 | 2 (0·03) | 0 | 0·03 (-0·01, 0·07) |
| Polymyalgia rheumatica | 1 (0·02) | 0 | 0·02 (-0·02, 0·06) | 1 (0·09) | 1 (0·12) | -0·04 (-0·30, 0·22) | 2 (0·03) | 1 (0·02) | 0·01 (-0·04, 0·06) |

| | | | | | | | | | |
|---|---|---|---|---|---|---|---|---|---|
| Rheumatoid arthritis | 1 (0·02) | 2 (0·05) | -0·04 (-0·13, 0·04) | 1 (0·09) | 1 (0·12) | 0 (-0·31, 0·30) | 2 (0·03) | 3 (0·06) | -0·04 (-0·12, 0·05) |
| Psoriatic arthropathy | 1 (0·02) | 0 | 0·02 (-0·02, 0·06) | 0 | 0 | 0 | 1 (0·01) | 0 | 0·02 (-0·02, 0·05) |
| Arthritis reactive | 0 | 1 (0·02) | -0·02 (-0·06, 0·02) | 0 | 0 | 0 | 0 | 1 (0·02) | -0·02 (-0·05, 0·02) |
| **Skin and subcutaneous tissue disorders** | 6 (0·09), (0·03, 0·21) | 2 (0·05), (0·01, 0·18) | 0·03 (-0·07, 0·14) | 0 | 0 | 0 | 6 (0·08), (0·03, 0·17) | 2 (0·04), (0·00, 0·15) | 0·03 (-0·06, 0·12) |
| Alopecia areata | 2 (0·03) | 0 | 0·03 (-0·01, 0·06) | 0 | 0 | 0 | 2 (0·03) | 0 | 0·02 (-0·01, 0·05) |
| Psoriasis | 2 (0·03) | 0 | 0·03 (-0·01, 0·06) | 0 | 0 | 0 | 2 (0·03) | 0 | 0·02 (-0·01, 0·05) |
| Erythema nodosum | 1 (0·02) | 0 | 0·01 (-0·01, 0·04) | 0 | 0 | 0 | 1 (0·01) | 0 | 0·01 (-0·01, 0·03) |
| Pemphigoid | 1 (0·02) | 0 | 0·02 (-0·02, 0·06) | 0 | 0 | 0 | 1 (0·01) | 0 | 0·02 (-0·02, 0·05) |
| Lichen planus | 0 | 1 (0·02) | -0·03 (-0·08, 0·03) | 0 | 0 | 0 | 0 | 1 (0·02) | -0·02 (-0·07, 0·02) |
| Lichenoid keratosis | 0 | 1 (0·02) | -0·03 (-0·08, 0·03) | 0 | 0 | 0 | 0 | 1 (0·02) | -0·02 (-0·07, 0·02) |
| **Endocrine disorders** | 3 (0·05), (0·01, 0·14) | 1 (0·02), (0·00, 0·14) | 0·01 (-0·06, 0·08) | 1 (0·09), (0·00, 0·49) | 0, (NA, 0·46) | 0·07 (-0·07, 0·21) | 4 (0·05), (0·01, 0·14) | 1 (0·02), (0·00, 0·11) | 0·02 (-0·04, 0·09) |
| Basedow's disease | 1 (0·02) | 0 | 0·01 (-0·01, 0·04) | 1 (0·09) | 0 | 0·07 (-0·07, 0·21) | 2 (0·03) | 0 | 0·02 (-0·01, 0·05) |
| Autoimmune thyroiditis | 1 (0·02) | 1 (0·02) | -0·01 (-0·08, 0·05) | 0 | 0 | 0 | 1 (0·01) | 1 (0·02) | -0·01 (-0·06, 0·04) |
| Hyperthyroidism | 1 (0·02) | 0 | 0·01 (-0·01, 0·04) | 0 | 0 | 0 | 1 (0·01) | 0 | 0·01 (-0·01, 0·03) |
| **Eye disorders** | 4 (0·06), (0·02, 0·16) | 1 (0·02), (0·00, 0·14) | 0·03 (-0·05, 0·10) | 0 | 0 | 0 | 4 (0·05), (0·01, 0·14) | 1 (0·02), (0·00, 0·11) | 0·02 (-0·04, 0·09) |
| Uveitis | 3 (0·05) | 1 (0·02) | 0·01 (-0·06, 0·08) | 0 | 0 | 0 | 3 (0·04) | 1 (0·02) | 0·01 (-0·05, 0·07) |
| Iridocyclitis | 1 (0·02) | 0 | 0·01 (-0·01, 0·04) | 0 | 0 | 0 | 1 (0·01) | 0 | 0·01 (-0·01, 0·03) |
| **Blood and lymphatic system disorders** | 2 (0·03), (0·00, 0·11) | 1 (0·02), (0·00, 0·14) | 0·01 (-0·07, 0·08) | 0, (NA, 0·33) | 1 (0·12), (0·00, 0·69) | -0·11 (-0·33, 0·11) | 2 (0·03), (0·00, 0·10) | 2 (0·04), (0·00, 0·15) | -0·01 (-0·08, 0·06) |
| Thrombocytopenia | 2 (0·03) | 1 (0·02) | 0·01 (-0·07, 0·08) | 0 | 1 (0·12) | -0·11 (-0·33, 0·11) | 2 (0·03) | 2 (0·04) | -0·01 (-0·08, 0·06) |
| **Gastrointestinal disorders** | 1 (0·02), (0·00, 0·09) | 2 (0·05), (0·01, 0·18) | -0·04 (-0·11, 0·04) | 1 (0·09), (0·00, 0·49) | 0, (NA, 0·46) | 0·07 (-0·07, 0·21) | 2 (0·03), (0·00, 0·10) | 2 (0·04), (0·00, 0·15) | -0·02 (-0·08, 0·05) |
| Colitis ulcerative | 0 | 0 | 0 | 1 (0·09) | 0 | 0·07 (-0·07, 0·21) | 1 (0·01) | 0 | 0·01 (-0·01, 0·03) |
| Crohn's disease | 1 (0·02) | 1 (0·02) | -0·01 (-0·06, 0·04) | 0 | 0 | 0 | 1 (0·01) | 1 (0·02) | -0·01 (-0·05, 0·03) |
| Coeliac disease | 0 | 1 (0·02) | -0·03 (-0·08, 0·03) | 0 | 0 | 0 | 0 | 1 (0·02) | -0·02 (-0·07, 0·02) |

| | | | | | | | | | |
|---|---|---|---|---|---|---|---|---|---|
| **Cardiac disorders** | 1 (0·02), (0·00, 0·09) | 0, (NA, 0·09) | 0·02 (-0·02, 0·06) | 0 | 0 | 0 | 1 (0·01), (0·00, 0·07) | 0, (NA, 0·08) | 0·02 (-0·02, 0·05) |
| Myocarditis | 1 (0·02) | 0 | 0·02 (-0·02, 0·06) | 0 | 0 | 0 | 1 (0·01) | 0 | 0·02 (-0·02, 0·05) |
| **Injury, poisoning and procedural complications** | 1 (0·02), (0·00, 0·09) | 0, (NA, 0·09) | 0·02 (-0·02, 0·06) | 0 | 0 | 0 | 1 (0·01), (0·00, 0·07) | 0, (NA, 0·08) | 0·02 (-0·02, 0·05) |
| Chilblains | 1 (0·02) | 0 | 0·02 (-0·02, 0·06) | 0 | 0 | 0 | 1 (0·01) | 0 | 0·02 (-0·02, 0·05) |
| **Investigations** | 1 (0·02), (0·00, 0·09) | 0, (NA, 0·09) | 0·01 (-0·01, 0·04) | 0 | 0 | 0 | 1 (0·01), (0·00, 0·07) | 0, (NA, 0·08) | 0·01 (-0·01, 0·03) |
| Heparin-induced thrombocytopenia test | 1 (0·02) | 0 | 0·01 (-0·01, 0·04) | 0 | 0 | 0 | 1 (0·01) | 0 | 0·01 (-0·01, 0·03) |

PIMMC=potential immune-mediated medical condition.
*Risk difference and its Confidence Intervals (CIs) are computed from Mantel-Haenszel Standardized Risk Estimates and 95% normal confidence limits with the stratification by study, while individual group statistics are not adjusted by strata. MedDRA version: 23·0 (2019nCoV-101 Part 1, 2019nCoV-101 Part 2, and 2019nCoV-501) and 23·1 (2019nCoV-301 and 2019nCoV-302)
